# Musical intervention to reduce stress during botulinum toxin injection for spasticity: protocol for a randomized controlled trial (MUSIBOT)

**DOI:** 10.1101/2025.06.13.25329602

**Authors:** Pierre Angelvy, Marina Badin, Mathilde Pelletier-Visa, Pascale Givron, Bruno Pereira, Emmanuel Coudeyre

## Abstract

**Introduction:** Botulinum toxin injections are a common treatment for managing spasticity resulting from central nervous system damage, including stroke, multiple sclerosis, and traumatic brain injury. However, the injections are associated with perceived pain, and many patients experience significant anticipatory stress regarding future sessions. The intensity of this stress varies among individuals.

Music therapy, particularly receptive musical interventions structured around a U-shaped sequence, promotes progressive relaxation through distinct musical phases. This method has demonstrated efficacy in reducing pain and anxiety across various clinical contexts, including chronic and acute pain, Alzheimer’s disease, fibromyalgia, and neurologically mediated pain. Given the painful nature of botulinum toxin injections, this study proposes the use of receptive music therapy to improve patient tolerance of the procedure. We hypothesize that receptive musical intervention can reduce injection-induced stress in adults undergoing botulinum toxin treatment. To our knowledge, no studies have specifically investigated the effect of music therapy on stress related to botulinum toxin injections.

We aim to conduct a prospective randomized (1:1) controlled trial to evaluate the impact of receptive music intervention on stress levels, measured via heart rate variability (HRV), during botulinum toxin injection sessions.

**Methods and analysis:** The primary objective is to assess the effect of receptive musical intervention during botulinum toxin injections on injection-induced stress, measured by HRV. Secondary objectives include evaluating the intervention’s effects on pain intensity and anxiety levels. Patient satisfaction following the music-assisted injection session will also be assessed. Additionally, the physician’s evaluation of the procedure and the patient’s perception of time during the session will be recorded.

**Ethics and dissemination:** All participants will provide written informed consent prior to enrollment. The study has received approval from the relevant institutional ethics committee (Comité de Protection des Personnes – ID: 25.00156.000468, Sud-Méditerranée IV, approved on 3 April 2025). Findings will be disseminated through peer-reviewed publications and presentations at scientific conferences.

**Strengths and limitations of this study:**

- This study will be the first to assess the effects of a receptive music intervention on stress during botulinum toxin injections for spasticity in adults.
- In addition to being non-invasive and non-pharmacological, music intervention offers multiple advantages, including minimal adverse effects, low cost, and easy accessibility and portability.
- The study design aims to achieve the highest level of evidence through a prospective, randomized, controlled approach.
- Stress is an abstract concept and there is no gold standardfor its evaluation; in this study, it will be assessed indirectly through heart rate variability.
- Receptivity to music intervention may vary between patients, potentially acting as a confounding factor.
- A habituation effect may occur in patients who have been exposed to this intervention over several years, potentially reducing observed stress levels.

## INTRODUCTION

Botulinum toxin (BT) injections are an effective treatment for spasticity caused by central neurological system lesions (1). and are widely used in people with conditions such as stroke, multiple sclerosis, spinal cord injury and traumatic brain injury. Botulinum toxin has been a reference treatment for focal spasticity since the recommendations of the SOFMER of 2009(2) . However, as the effect of the botulinum toxin lasts approximately 3 months, the injections must be repeated several times per year. These injections are often perceived as painful. As well as the skin break in and the injection of the product, the technique used to guide the injections may cause pain, particularly electrical stimulation. Some injection sites are more painful than others, such as the palms of the hands and soles of the feet (3). Injection tolerance varies across individuals, but most experience significant levels of stress during BT injections.

Stress-related body reactions are induced by the autonomic nervous system. Stress can be objectively assessed using biomarkers such as heart rate variability (HRV)(4). Heart rate measurement is non-invasive, painless, easy to perform, and reproducible (5). It reflects the cardiovascular response to regulatory impulses affecting heart rhythm (6). Heart rate variability is a reliable indicator of autonomic nervous system activity (7), and numerous studies have employed HRV to estimate mental stress (8–11).

Since ancient times, music has been used to treat various ailments (12). Reintroduced for therapeutic purposes in the 1960s, musical interventions have since been employed to alleviate pain and distress in patients with a range of medical conditions, particularly in the treatment of chronic and acute pain (13,14). A recent meta-analysis found that musical interventions had positive effects on pain intensity, emotional distress, the use of anesthetics and opioid and non-opioid agents, heart rate, systolic and diastolic blood pressure, and respiratory rate (15). However, contradictory results have been reported, likely due to the wide variability in musical interventions across different studies, which vary in terms of duration, frequency, style, genre, preparation, selection, and method of implementation (15). To address these inconsistencies, recommendations have been formulated for the use of musical interventions in clinical practice (16,17). In line with these guidelines, a standardized musical intervention based on a web application was developed at the University Hospital Center of Montpellier. This intervention follows a U-shaped composition sequence lasting 20 to 60 minutes, structured in several stages designed to gradually guide the patient into relaxation according to the U-technique (18). The application allows for controlled use by the patient and/or caregiver at the bedside via a tablet. Based on the U-sequence, the Music Care program provides access to a range of music tracks pre-recorded by professionals in music therapy, musicians (19).

Several studies have focused on stress management, pain reduction, anxiety relief, and improving quality of life in patients with various pathologies and painful conditions, including cataract surgery (20,21), migraine treatment (22), coronary angiography (23), and cancer treatment (24).

To our knowledge, no clinical trials have examined the use of music interventions in conjunction with BT injections for the treatment of spasticity. The aim of this study is to investigate the effect of a music intervention session during BT injections on injection-induced stress. Our research team previously investigated another analgesic technique using virtual reality during BT injections in a separate study (25).

We hypothesize that a receptive music intervention can reduce the stress caused by BT injections in adults.

## METHODS AND ANALYSIS

### Study design and setting

We will conduct a prospective, interventional, randomized controlled trial in the Physical and Rehabilitation Medicine (PRM) department of the Clermont Ferrand University Hospital, France.

Participants will be assigned to one of two groups: control or experimental. The control group will follow a waiting-list design, meaning participants in this group will receive the music intervention during their second injection. Each patient will serve as their own control, undergoing a first standard injection session without the music therapy device but with the heart rate sensor in place.

This study protocol is reported in accordance with the Standard Protocol Items: Recommendations for Interventional Trials (SPIRIT) guidelines, presented in the appendix (26). The study results will be reported following the CONSORT Statement for non-pharmacologic trials (27). Interventions are described in detail according to the Template for Intervention Description and Replication (TIDieR) checklist (28). The provisional study schedule is shown in **Figure 1**.

**Figure 1.**
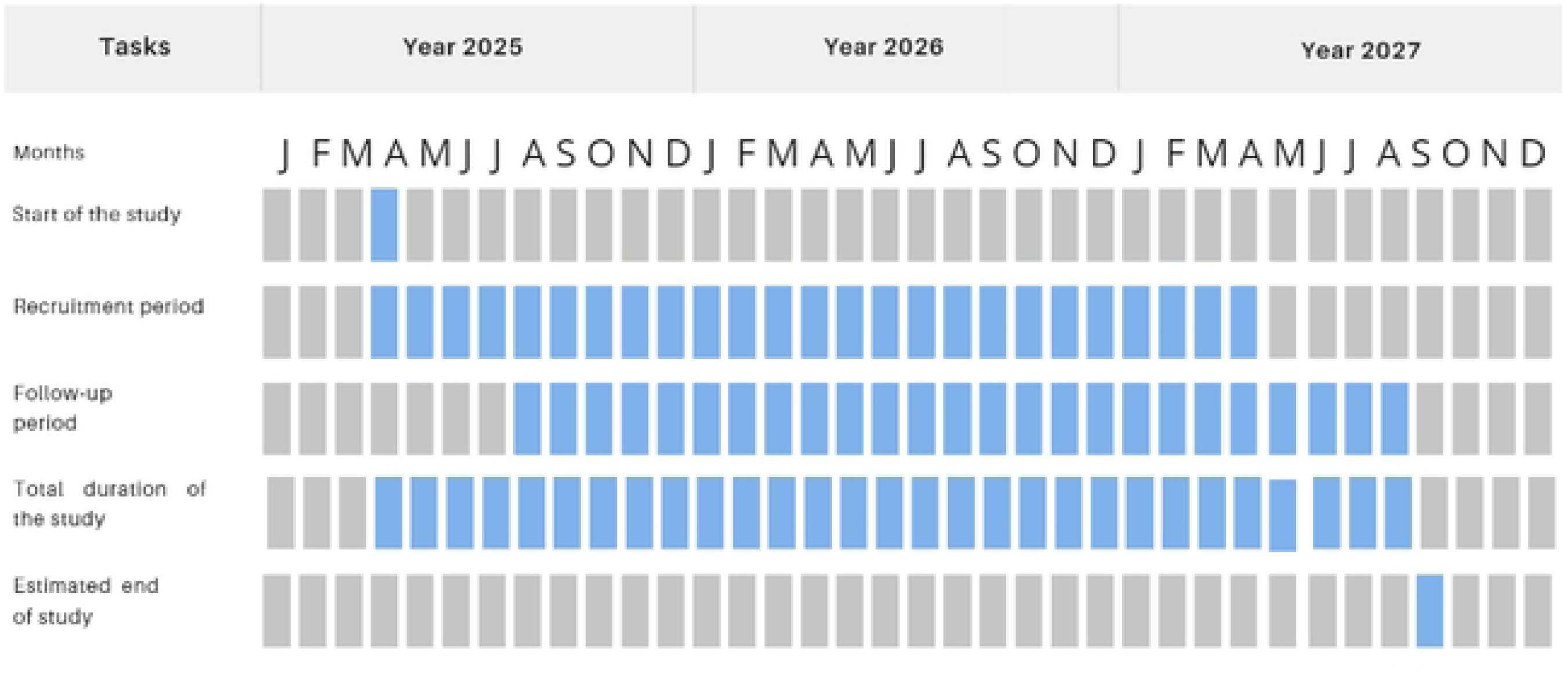
Provisional study schedule of MUSIBOT

### Participants

Eligible participants for this study are adult patients, male or female, with neurological related spasticity, such as that resulting from multiple sclerosis, stroke, or traumatic brain injury, who are candidates for BT injection treatments. Participants may have undergone previous BT injections. Participants must have a known history of pain and/or anxiety and be capable of providing informed consent to take part in the research. Additionally, they must be affiliated with the French Social Security insurance scheme.

Patients with contraindications to musical interventions, such as severe hearing impairment, unstable psychotic disorders, or a history of auditory trauma will be excluded. People with major cognitive disorders and any other medical condition deemed incompatible with the study by the investigator will result in exclusion. Patients requiring sedation with nitrous oxide during BT injection sessions will not be eligible. Additionally, medications or with medical conditions that may interfere with HRV, such as beta-blockers, antiarrhythmics, anxiolytics, benzodiazepines, antihypertensives, and calcium channel blockers, will be considered exclusion criteria. Pregnant or breastfeeding women, as well as individuals who decline to participate, will also be excluded.

### Recruitment

Participants will be recruited from the PRM departments of Clermont-Ferrand University Hospital in France. The CRA, in collaboration with the study investigators (physicians), will verify the eligibility criteria of patients followed in the department. Information about the study, along with an information sheet, will be provided to eligible patients 1 to 3 months before the inclusion visit, allowing them sufficient time to consider participation. For patients under protective measures, their guardian or curator must be present to review the information sheet. During the inclusion visit, individuals and/or their relatives will inform the team of their decision to participate or not. If both the patient and their relatives agree, the written consent form will be signed by the patient and/or their legal guardian or curator, as well as by the study investigator.

### Randomization

Participants will be randomized (1:1) into the experimental or control group during the first consultation. Block randomization with varying block sizes will be used. The randomization will be carried out by the CRA using RedCap (Version 8.2.50).

### Interventions

All participants will undergo a first injection session without the Music Care application, wearing a Polar H10 heart rate monitor. The second session will be performed 3 – 4 months later as is standard practice for BT injection. All participants will wear the Polar H10 heart rate monitor and they will use the Music Care application or not, according to group allocation.

### Intervention group

The Music Care application is a receptive musical intervention in which patients select music based on their personal preferences from a variety of styles available on a tablet. A single, individualized music session is administered during the BT injection session. The music is played through headphones, in a calm environment conducive to relaxation. The standard musical sequence, lasting 20 to 60 minutes, is structured in several phases designed to gradually induce a state of relaxation, following the “U-sequence” method. This U-shaped sequence involves a progressive decrease in musical tempo, orchestral intensity, frequencies, and volume (the downward phase of the U) until a period of maximum relaxation (the bottom of the U) is reached. This is followed by a gradual recovery phase (the upward segment of the U)(18). The U-sequence method is illustrated in Figure 2. All musical sequences, designed according to this method, were recorded by Music Care (Paris, France).

**Figure 2.**
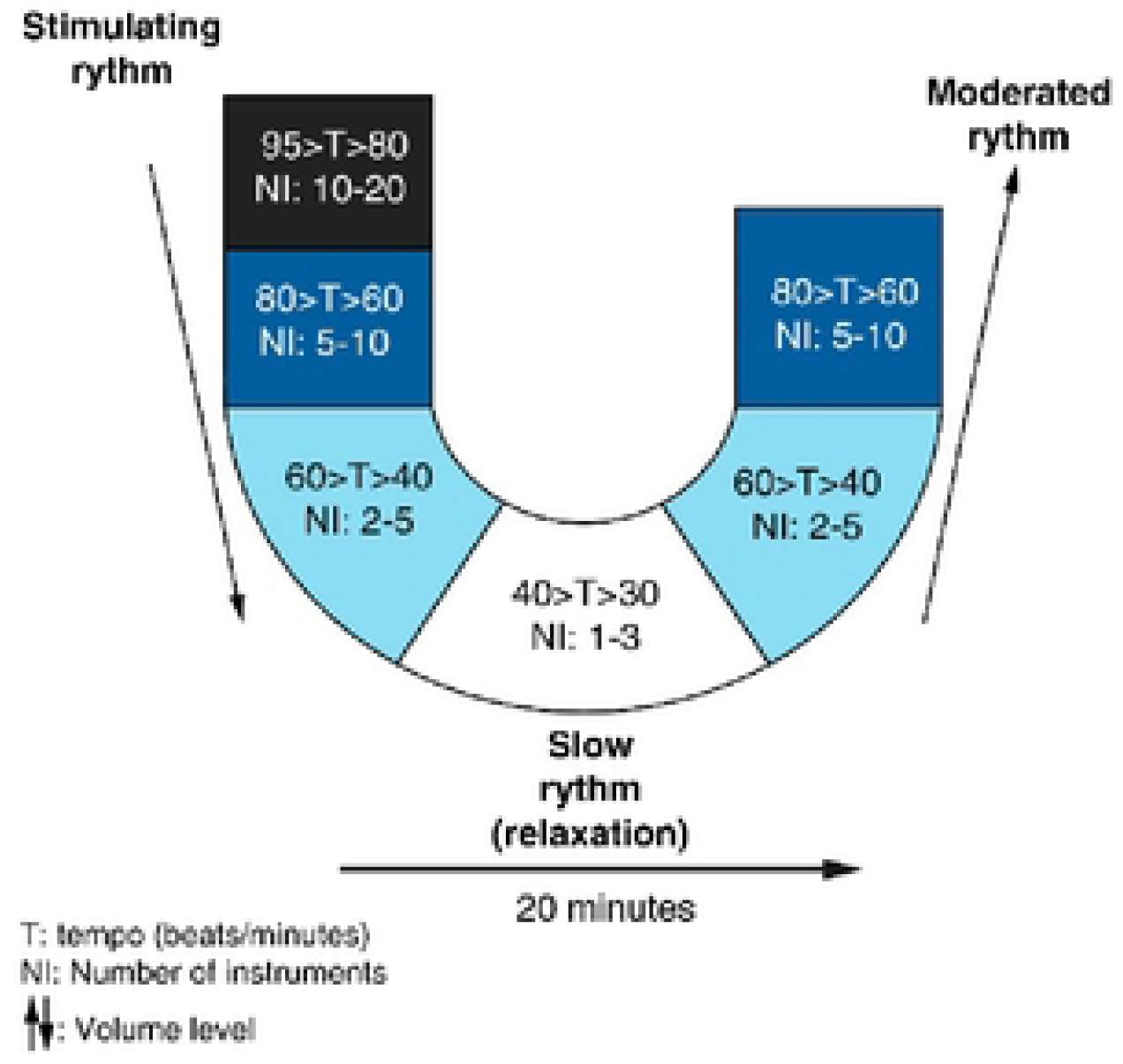
The U sequence method

No pain medication beyond the patient’s usual prescription will be permitted during the procedure.

During the injections, the same protocol will be consistently applied for each participant: either ultrasound or electrostimulation will be used to guide the injection, a MYOBOT needle will be used, and ice analgesia fwill be administered for palmar and plantar injections.

### Control group

The control group will not receive any specific intervention, apart from the use of the Polar H10 heart rate monitor. Participants in this group will undergo BT injections as routinely performed in the department, and they will undergo the same assessments as the group receiving the music intervention during both injection sessions.

### Primary outcome

The primary outcome is the effect of the music intervention on stress, assessed through HRV during the first injection visit. This will be estimated by the variation in HRV measured before and during the BT injection session. Data will be collected using a Polar H10 heart rate monitor, exported to the Elite HRV application, and analyzed with KUBIOS software.

### Secondary outcomes

The secondary outcomes include several aspects of the patient’s experience during the BT injection sessions. First, patient stress will be assessed again via HRV measured before and during the second injection visit. Pain intensity will be recorded during both the first and second injection visits using a numeric rating scale from 0 (no pain) to 10 (worst pain imaginable), immediately after each session. Anxiety levels will be evaluated using the State-Trait Anxiety Inventory Form Y-State (STAI-Y1)(29), a self-report questionnaire administered before and after both injection visits. The patient’s perception of time during the session will also be measured by comparing their subjective experience of time to the actual injection duration, with data collected at the end of each session. Patient satisfaction with the intervention will be assessed using a visual analog scale from 0 (very dissatisfied) to 10 (very satisfied), following both injection visits. Finally, the quality of the conditions for administering the BT injections will be rated by the physician immediately after each session, using a numeric scale from 0 (very poor condition) to 10 (very good condition).

## STATISTICAL CONSIDERATIONS

### Sample size estimation

To evaluate the effect of a music intervention session during BT injections on injection-induced stress, 35 patients per group will be required to detect an absolute difference in HRV of at least 1.8, with a standard deviation of 2.3. This calculation assumes a two-sided type I error of 5% and a statistical power of 90%. The expected difference and standard deviation are based on findings from a previous study conducted by our team (Clinical Trials NCT05364203). To account for potential dropouts or missing data, a total of 40 patients per group is proposed.

### Statistical analysis

All analyses will be performed using Stata software version 15 (StataCorp, College Station, TX, USA). Continuous variables will be presented as mean ± standard deviation or as median with interquartile range, depending on the data distribution. The assumption of normality will be assessed using the Shapiro–Wilk test.

At Injection Visit #1, the randomized groups will be compared in terms of eligibility compliance, epidemiological and clinical characteristics, and treatments. Baseline comparability between groups will be assessed using key participant characteristics and potential factors associated with the primary endpoint. Any observed imbalances will be interpreted based on clinical relevance rather than statistical significance alone. Protocol deviations, including their nature and the number of patients affected, will be documented along with the reasons for withdrawal. The number of enrolled patients and the inclusion flowchart will be reported by group.

Statistical analyses will be conducted primarily on the intention-to-treat population, including all participants except those who withdraw consent for data use. To minimize bias from informative missingness, appropriate imputation methods—such as multiple imputation—may be applied, depending on the extent and nature of the missing data, particularly for the primary and secondary endpoints. A secondary per-protocol analysis will be conducted, excluding participants who were not managed according to their assigned group or who had major protocol deviations. The type I error will be set at 5% (two-sided).

The primary objective is to assess the effect of exposure to a music intervention session during Injection Visit #1 on injection-induced stress, as measured by HRV. The primary analysis will compare the change in the low-frequency to high-frequency ratio (LF/HF ratio) of HRV between the randomized groups using Student’s t-test, or the Mann–Whitney U test if normality assumptions are not met. Equality of variances will be assessed using the Fisher–Snedecor test. Results will be reported as effect sizes with 95% confidence intervals. A multivariate analysis, such as multiple linear regression, will then be conducted to incorporate covariates identified through univariate analysis and clinical relevance (e.g., patient age and sex). Multicollinearity will be evaluated using Farrar and Glauber’s test and variance inflation factor indicators. Results will include regression coefficients, effect sizes, and 95% confidence intervals. Subgroup analyses will explore the primary outcome by sex and age, examining group-by-subgroup interactions, with results reported in the same manner.

Secondary analyses will assess the effect of exposure to a music intervention session during BT injections at Injection Visit #1 on injection-induced stress (excluding the LF/HF ratio), pain, anxiety, patient satisfaction, and the physician-rated impact of the music intervention on the procedure. These analyses will follow the same statistical approach as used for the primary endpoint. To evaluate the effect of repeated exposure to the music intervention at Injection Visit #2, changes in stress, pain, anxiety, patient satisfaction, and physician-rated procedural impact will be analyzed using mixed-effects models. Longitudinal repeated measures will be modeled using random-effects models that account for the fixed effects of group, time, and their interaction, as well as inter- and intra-subject variability. More specifically, a constrained longitudinal data analysis mixed-effects linear model will be used for continuous outcomes, incorporating both baseline and follow-up values, with baseline group differences constrained to zero. This approach allows estimation of changes over time adjusted for baseline, with between-group differences captured via the group-by-time interaction. Random intercept and slope effects at the patient level will be included, and model parameters will be estimated using restricted maximum likelihood. Within-group analyses will also be conducted. For categorical outcomes, generalized linear mixed-effects models with a logit link and a constrained longitudinal data analysis framework will be applied to estimate odds ratios at follow-up, adjusted for baseline, with random effects modeled similarly. Non-repeated data will be analyzed using Student’s t-test or the Mann–Whitney U test for continuous variables (e.g., patient satisfaction), depending on normality and variance assumptions, and the chi-square test or Fisher’s exact test for categorical variables.

## ETHICS AND DISSEMINATION

The study protocol was approved by the Medical Ethics Committee of Sud-Méditerranée IV and the Agence Nationale de Sécurité des Médicaments et des Produits de Santé (ANSM) the French National Agency for Medicine and Health Product Safety on March 4, 2025 (No. 25.00156.000468). All participants will provide written informed consent prior to enrollment, and the study will be conducted in accordance with the principles outlined in the Declaration of Helsinki.

Participants will be informed of their right to withdraw from the study at any time, in accordance with Good Clinical Practice guidelines and the French regulatory framework. Premature withdrawal may also occur due to intercurrent illness, death, significant protocol deviations, or loss to follow-up. Any adverse events reported during the study will be promptly communicated to the principal investigator to ensure timely action in safeguarding participants’ health. Data confidentiality will be strictly maintained, with robust measures in place to protect participants’ identities.

Any protocol modifications affecting study conduct, participant safety, or potential benefits will require formal approval through an amendment process by the CPP Sud-Méditerranée IV to ensure continued compliance with ethical standards.

The study findings will be disseminated through peer-reviewed publications and presentations at scientific conferences.

## DISCUSSION

This study aims to assess the effects of a receptive musical intervention on stress during BT injections for spasticity in adults. The anticipated benefit for patients is improved tolerance of the injection procedure.

Music intervention offers several advantages: it is non-invasive, non-pharmacological, low-cost, and easily accessible and portable. By helping to isolate the patient from the clinical environment, it enables the individual to focus on the musical experience and become distracted from the unpleasant stimuli of a stressful medical setting (30).

Although stress is an abstract concept, it can be quantified through HRV, which is sensitive to sympathomimetic influences and requires highly standardized conditions. In general, HRV is a reliable indicator of autonomic nervous system activity (31), and many previous studies have used it to estimate mental stress(8–11).

One limitation worth noting is individual variability in receptivity to music therapy. Responses may differ from one patient to another, potentially introducing a confounding factor. However, the sample size was calculated to account for this variability. Another possible limitation is the habituation effect—patients who have been receiving this therapy for several years may experience less stress overall, which could influence the study outcomes.

## Data Availability

Data will be disclosed only following prior mutual agreement between the investigator and the sponsor.

## Data monitoring and auditing

No monitoring is planned for this study.

## Data access and transparency

Access to the final trial data will be strictly controlled to ensure participant confidentiality and maintain research integrity. The principal investigator (MB) and the designated biostatistician (BP) will have full access to the data for in-depth analysis and accurate interpretation. To promote transparency and scientific collaboration, anonymized data will also be made available upon request to researchers affiliated with recognized research institutions. This approach supports reproducibility and enables valuable secondary analyses.

## Authors’ contributions

The guarantor of the study is the corresponding author, PA. The study was conceived and designed by PA, MPV, MB, and EC. The original protocol was drafted by PA, MPV, MB, BP, and EC. Study coordination was carried out by MB and MPV. Data acquisition was performed by PA, MB, and MPV. The statistical analysis plan was designed by BP and EC. The present manuscript was drafted by PA, MPV, MB, BP, and EC, and all authors provided final approval of the submitted version.

## Communication policy

Data will be disclosed only following prior mutual agreement between the investigator and the sponsor. The study results will be communicated and published in accordance with international publication guidelines (www.icmje.org). Authors affiliated with Clermont-Ferrand University Hospital will be listed as the first and last authors of the primary publication. The study will be registered on the ClinicalTrials.gov website.

## REFERENCES

1. Santamato A, Cinone N, Panza F, Letizia S, Santoro L, Lozupone M, et al. Botulinum Toxin Type A for the Treatment of Lower Limb Spasticity after Stroke. Drugs. févr 2019;79(2):143–60.

2. sofmer_Reco-Spasticite.pdf [Internet]. [cité 3 juin 2025]. Disponible sur: https://www.sofmer.com/download/sofmer_Reco-Spasticite.pdf

3. Mathevon L, Bonan I, Barnais JL, Boyer F, Dinomais M. Adjunct therapies to improve outcomes after botulinum toxin injection in children: A systematic review. Ann Phys Rehabil Med. juill 2019;62(4):283–90.

4. Dutheil F, Boudet G, Perrier C, Lac G, Ouchchane L, Chamoux A, et al. JOBSTRESS study: Comparison of heart rate variability in emergency physicians working a 24-hour shift or a 14-hour night shift — A randomized trial. Int J Cardiol. juill 2012;158(2):322–5.

5. Heart rate variability: standards of measurement, physiological interpretation and clinical use. Task Force of the European Society of Cardiology and the North American Society of Pacing and Electrophysiology. Circulation. 1 mars 1996;93(5):1043–65.

6. Kim HG, Cheon EJ, Bai DS, Lee YH, Koo BH. Stress and Heart Rate Variability: A Meta-Analysis and Review of the Literature. Psychiatry Investig. mars 2018;15(3):235–45.

7. Kumar M, Weippert M, Vilbrandt R, Kreuzfeld S, Stoll R. Fuzzy Evaluation of Heart Rate Signals for Mental Stress Assessment. IEEE Trans Fuzzy Syst [Internet]. 1 oct 2007 [cité 3 juin 2025]; Disponible sur: https://dl.acm.org/doi/10.1109/TFUZZ.2006.889825

8. Yashima K, Sasaki T, Kageyama Y, Odagaki M, Hosaka H. Application of wavelet analysis to the plethysmogram for the evaluation of mental stress. In IEEE; 2005 [cité 22 déc 2021]. p. 2781–4. Disponible sur: http://ieeexplore.ieee.org/document/1617049/

9. Sato N, Miyake S. Cardiovascular Reactivity to Mental Stress: Relationship with Menstrual Cycle and Gender. J Physiol Anthropol Appl Human Sci. 2004;23(6):215–23.

10. Orsila R, Virtanen M, Luukkaala T, Tarvainen M, Karjalainen P, Viik J, et al. Perceived Mental Stress and Reactions in Heart Rate Variability—A Pilot Study Among Employees of an Electronics Company. Int J Occup Saf Ergon. janv 2008;14(3):275–83.

11. Melillo P, Bracale M, Pecchia L. Nonlinear Heart Rate Variability features for real-life stress detection. Case study: students under stress due to university examination. Biomed Eng OnLine. 2011;10(1):96.

12. Marwick C. Music Therapists Chime In With Data on Medical Results. JAMA. 9 févr 2000;283(6):731–3.

13. Brandes V, Terris DD, Fischer C, Loerbroks A, Jarczok MN, Ottowitz G, et al. Receptive music therapy for the treatment of depression: a proof-of-concept study and prospective controlled clinical trial of efficacy. Psychother Psychosom. 2010;79(5):321–2.

14. Dileo C. Effects of music and music therapy on medical patients: a meta-analysis of the research and implications for the future. J Soc Integr Oncol. 2006;4(2):67–70.

15. Lee JH. The Effects of Music on Pain: A Meta-Analysis. J Music Ther. 1 déc 2016;53(4):430–77.

16. Kamioka H, Tsutani K, Yamada M, Park H, Okuizumi H, Tsuruoka K, et al. Effectiveness of music therapy: a summary of systematic reviews based on randomized controlled trials of music interventions. Patient Prefer Adherence. 2014;8:727–54.

17. Nilsson U. The anxiety- and pain-reducing effects of music interventions: a systematic review. AORN J. avr 2008;87(4):780–807.

18. Guétin S, Touchon J. Musique et douleur : la séquence en « U », une solution thérapeutique standardisée et validée. Douleur Analgésie [Internet]. 11 juill 2017 [cité 18 mars 2024]; Disponible sur: 10.1007/s11724-017-0507-2

19. Guétin S, Giniès P, Siou DKA, Picot MC, Pommié C, Guldner E, et al. The Effects of Music Intervention in the Management of Chronic Pain: A Single-Blind, Randomized, Controlled Trial. Clin J Pain. mai 2012;28(4):329.

20. Guerrier G, Abdoul H, Jilet L, Rothschild PR, Baillard C. Efficacy of a Web App-Based Music Intervention During Cataract Surgery: A Randomized Clinical Trial. JAMA Ophthalmol. 1 sept 2021;139(9):1007–13.

21. Guerrier G, Abdoul H, Jilet L, Rothschild PR, Levy J, Rondet S, et al. Musical intervention reduces anxiety-related hypertensive events during cataract surgery: A randomized controlled trial. Perioper Care Oper Room Manag. sept 2020;20:100126.

22. Parlongue G, Cerdan EV, Koenig J, Williams DP. Smartphone based music intervention in the treatment of episodic migraine headaches - A pilot trial. Complement Ther Med. déc 2021;63:102779.

23. Guétin S, Brun L, Deniaud M, Clerc JM, Thayer JF, Koenig J. Smartphone-based Music Listening to Reduce Pain and Anxiety Before Coronarography: A Focus on Sex Differences. Altern Ther Health Med. juill 2016;22(4):60–3.

24. Krishnaswamy P, Nair S. Effect of Music Therapy on Pain and Anxiety Levels of Cancer Patients: A Pilot Study. Indian J Palliat Care. 2016;22(3):307–11.

25. Bougeard M, Hauret I, Pelletier-Visa M, Plan-Paquet A, Givron P, Badin M, et al. Use of immersive virtual reality for stress reduction during botulinum toxin injection for spasticity (RVTOX): a study protocol of a randomised control trial. BMJ Open. 30 oct 2023;13(10):e066726.

26. Chan AW, Boutron I, Hopewell S, Moher D, Schulz KF, Collins GS, et al. SPIRIT 2025 statement: updated guideline for protocols of randomized trials. Nat Med. 29 avr 2025;1–9.

27. Boutron I, Altman DG, Moher D, Schulz KF, Ravaud P, CONSORT NPT Group. CONSORT Statement for Randomized Trials of Nonpharmacologic Treatments: A 2017 Update and a CONSORT Extension for Nonpharmacologic Trial Abstracts. Ann Intern Med. 4 juill 2017;167(1):40–7.

28. van Vliet P, Hunter SM, Donaldson C, Pomeroy V. Using the TIDieR Checklist to Standardize the Description of a Functional Strength Training Intervention for the Upper Limb After Stroke. J Neurol Phys Ther. juill 2016;40(3):203–8.

29. Saviola F, Pappaianni E, Monti A, Grecucci A, Jovicich J, De Pisapia N. Trait and state anxiety are mapped differently in the human brain. Sci Rep. 6 juill 2020;10(1):11112.

30. Lombard M, Ditton T. At the Heart of It All: The Concept of Presence. J Comput-Mediat Commun. 23 juin 2006;3(2):0–0.

31. Kumar M, Weippert M, Vilbrandt R, Kreuzfeld S, Stoll R. Fuzzy Evaluation of Heart Rate Signals for Mental Stress Assessment. IEEE Trans Fuzzy Syst. oct 2007;15(5):791–808.

